# A Metabolomics Approach for Early Prediction of Vincristine-Induced Peripheral Neuropathy

**DOI:** 10.1101/19013078

**Authors:** Parul Verma, Jayachandran Devaraj, Jodi L. Skiles, Tammy Sajdyk, Richard H. Ho, Raymond Hutchinson, Elizabeth Wells, Lang Li, Jamie Renbarger, Bruce Cooper, Doraiswami Ramkrishna

## Abstract

Vincristine is a core chemotherapeutic drug administered to pediatric acute lymphoblastic leukemia patients. Despite its efficacy in treating leukemia, it can lead to severe peripheral neuropathy in a sub-group of the patients. Peripheral neuropathy is a debilitating and painful side-effect that can severely impact an individual’s quality of life. Currently, there are no established predictors of peripheral neuropathy incidence during the early stage of chemotherapeutic treatment. As a result, patients who are not susceptible to peripheral neuropathy may receive sub-therapeutic treatment due to an empirical upper cap on the dose, while others may experience severe neuropathy at the same dose. Contrary to previous genomics based approaches, we employed a metabolomics approach to identify small sets of metabolites that can be used to predict a patient’s susceptibility to peripheral neuropathy at different time points during the treatment. Using those identified metabolites, we developed a novel strategy to predict peripheral neuropathy and subsequently adjust the vincristine dose accordingly. In accordance with this novel strategy, we created a free user-friendly tool, *VIPNp*, for physicians to easily implement our prediction strategy. Our results showed that focusing on metabolites, which encompasses both genotypic and phenotypic variations, can enable early prediction of peripheral neuropathy in pediatric leukemia patients.

## Introduction

Acute lymphoblastic leukemia (ALL) is the most common cancer among children, accounting for approximately 26% of all pediatric cancers in the USA [1]. Although the 5-year survival rate of pediatric leukemia patients is as high as 86%, the side-effects of the treatment can severely impact the quality of life of survivors [2]. In particular, vincristine, a core chemotherapeutic drug administrated as part of the ALL therapy that has been in use for more than 50 years, has a dose-limiting toxicity: peripheral neuropathy. Vincristine-induced peripheral neuropathy (VIPN) is characterized primarily by numbness, tingling, and a painful sensation felt in the hands and feet, muscle weakness, and constipation due to its effect on the sensory, motor, and autonomic nerves [3–5]. In some instances, VIPN can be prolonged and may last even after discontinuation of the treatment, impairing patients’ motor skills [6–9] which results in limitation of their daily life activities for many years after completion of therapy [10]. While VIPN is severe in a subpopulation of patients, another subpopulation experiences negligible neuropathy. Currently, there are neither established ways of predicting susceptibility to VIPN in patients, nor ways to treat it effectively, resulting in a suboptimal management for both the cohorts. Predicting VIPN susceptibility in patients will enable better dosage decision making tools for physicians and in turn may improve the quality of life of these patients.

Several researchers have studied the association between genomics and VIPN incidence; however, the majority of the results have been controversial. Most of the studied differences are based on race, CYP3A5, ABC transporter, and, more recently, CEP72 expression. While some studies have reported a significant association between race and VIPN incidence [11–14], others could not confirm those results [5, 15]. CYP3A5 metabolizes vincristine and some studies have reported a significant association between CYP3A5 expression and VIPN [16, 17, 13]; however, one study could not establish such an association [18]. Similarly, there is evidence for association between the ABCB1 transporter and VIPN [19, 20]; and there also exists evidence to the contrary [18]. Additional details on the associations and non-associations found between VIPN and other variables is addressed by Velde et al. [4] and Mora et al. [3]. Correlations between pharmacokinetics or other patient-specific co-variates and VIPN have also been evasive due to a large variability in the available interpatient and intrapatient data, making interpretation of these pharmacokinetic studies difficult [21–25]. Recently, significant correlation was discovered of a SNP in the promoter region of the CEP72 gene with VIPN, during the maintenance phase of the treatment. CEP72 expression was associated with VIPN in human neurons derived from human induced pluripotent stem cells, as well [12]. While some studies have been able to reproduce this finding during the late phase of the treatment [26, 27], this association was not found in the earlier phase of therapy [28, 29]. The literature clearly shows that an established predictor for the early stage of the treatment is still lacking.

In this paper, we take a detour from the previous genomics and pharmacokinetics related studies and employ a novel approach using metabolomics to predict VIPN at an earlier stage of the treatment. The need to go beyond genomics has been raised before [30], and, specifically, the role of metabolomics in predicting drug response has garnered more attention [31, 32] in recent years. The recent interest in pairing metabolomics and drug response is a result of phenotypic variations that can be captured at the downstream metabolite level. These phenotypic variations arise due to differences in environment, lifestyle, stochasticity in biochemical reactions, etc., and may be as important as, if not more important than, the upstream genomics to predict drug response [30]. Information transfer for expression of a disease outcome takes place at various stages, starting from transcription of a gene to mRNA, then translation to a protein, and, finally, to the synthesis of metabolites. Variations at any stage may induce a different response to any drug. Because of this, drug response of a homogeneous population of patients with a similar genome may vary. Moreover, variations at the genetic level may not get transferred to the downstream metabolite level because of the robustness of the metabolic pathways. Therefore, a metabolomics approach is employed to find metabolites that can accurately predict VIPN in a cohort of pediatric patients who underwent ALL treatment, with vincristine as the core chemotherapy drug. We determined small sets of metabolites that can accurately predict VIPN at different stages of the treatment and used them to develop a strategy to identify patients that are highly susceptible to VIPN. Using this strategy, we developed a free user-friendly tool, *VIPNp* [33, 34], for physicians to apply our models for prediction of VIPN susceptibility at different stages of a patient’s chemotherapy treatment.

## Results

In this section, we elaborate on our workflow and the results. We categorized the neuropathy data obtained from physician evaluations of VIPN severity experienced by patients. We then performed hierarchical clustering to differentiate between metabolite profiles measured at both early and late time points of the treatment process. Based on their metabolite profiles measured at different time points, we found a small subset of biomarker metabolites which was used to build a predictive model of VIPN susceptibility. We were able to identify the molecular structure of some of the biomarkers and then performed pathway analysis.

### Pediatric ALL patient data description

A total of 36 patients’ data was collected. These were pediatric patients treated with AALL0932 protocol for B-ALL enrolled in a study from 2010 to 2014 (see Methods for details). Vincristine was part of the core chemotherapeutic treatment. All subjects were phenotyped for peripheral neuropathy using the Total Neuropathy Score – Pediatric Vincristine (TNS©-PV) [35]. Twenty-four of the 36 patients (67%) were phenotyped as patients experiencing high neuropathy (HN), while the remaining 12 patients (33%) were classified as patients experiencing low neuropathy (LN) after the treatment completion. Patient demographics are described in Table 1. Non-fasting blood samples for these patients were collected prior to administration of drugs at three time points of the treatment: during the induction phase (day 8 and day 29) and the consolidation phase (around 6 months).

**Table 1:** Patient characteristics were defined according to gender, age, and body mass index (BMI). For the day 8 treatment time point, 8 overall low neuropathy (LN) samples were available, while for day 29 and month 6 time points, 12 LN samples were available. 24 overall high neuropathy (HN) samples were available at all the three time points. Age and BMI correspond to that during the start of the treatment. Here, “HN” implies that the patient had a Total Neuropathy Score – Pediatric Vincristine (TNS©-PV) greater than 8 at least once, and “LN” implies that the patient had a TNS©-PV less than 3 throughout the treatment. SD: standard deviation.

|  | Day 8 |  | Day 29 and Month 6 |  |
| --- | --- | --- | --- | --- |
|  | HN | LN | HN | LN |
| Total | 24 | 8 | 24 | 12 |
| Females/Males | 11/13 | 4/4 | 11/13 | 5/7 |
| Age, in years (Mean, SD) | 9.6,4.8 | 4.3,2.4 | 9.6,4.8 | 4.4,2.6 |
| BMI, in Kg/m<sup>2</sup> (Mean, SD) | 24.4,12.4 | 14.5,2.6 | 24.4,12.4 | 15.1,2.8 |

Patients were assessed regularly using the TNS©-PV scoring scheme throughout the 2-3 years of treatment. A score less than 3 was considered of low intensity and a score greater than 8 was considered of high intensity. A patient was classified as LN if the TNS©-PV intensity remained low throughout the entire duration of the treatment which lasted 2-3 years, regardless of the drug amount. A patient was classified as HN if the TNS©-PV intensity was high at any time point during the treatment. This classification scheme matched the course of treatment for patients experiencing severe neuropathy. A patient who presents a score of 8 or higher at least once indicates susceptibility for severe neuropathy. The degree of neuropathy may improve during later phases of treatment when the drug is given less frequently or after dose modifications have been made for severe VIPN. However, these adjustments do not make a patient less susceptible to neuropathy in the future; if a patient was susceptible to high neuropathy even once, we classified the patient as HN. As a result of this classification system, patients who never had a TNS©-PV greater than 8 but had a TNS©-PV greater than 3 at some point (medium intensity) were not considered for the purposes of this study. This classification, as well as, threshold scores of 3 and 8 TNS©-PV were delineated to ensure a clear demarcation between patients who were susceptible to severe neuropathy and those who experienced negligible neuropathy.

Figure 1 shows how the neuropathy score changed during the course of the treatment time points considered and how it compared to the overall neuropathy susceptibility of a patient. At day 8, most of the patients had a low TNS©-PV intensity (TNS©-PV less than 3) while the incidence of high neuropathy (TNS©-PV greater than 8) increased with time. However, one patient had low TNS©-PV at day 8, high TNS©-PV at day 29, and low again at the month 6 time point. Figure 1 shows that by 6 months, 17 out of 24 HN (TNS©-PV greater than 8 at least once during the treatment) patients had already experienced high neuropathy, indicating a need for dosage adjustment earlier in the treatment process. Consequently, this data shows it is imperative to find biomarkers that enable the prediction of overall neuropathy susceptibility during the early stage of the treatment. The focus of this study is on finding metabolites that can differentiate between HN and LN during these time points.

**Figure 1:**
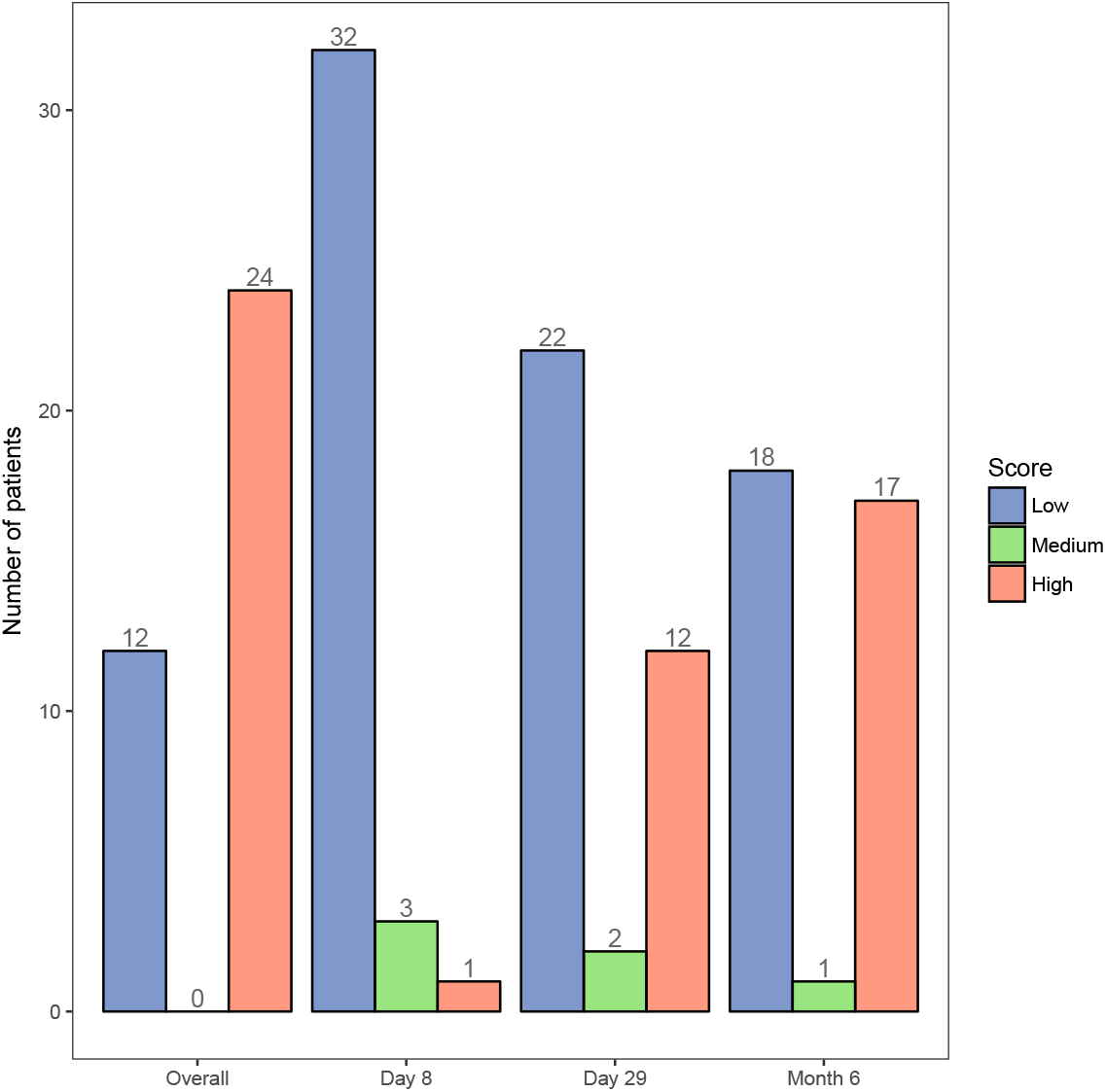
A bar plot showing distribution of the neuropathy score of patients over time. A TNS©-PV less than 3 corresponds to low, a score between 3 and 8 corresponds to medium, and a score above 8 corresponds to high. The first group shows the overall susceptibility of patients to neuropathy (LN versus HN), based on regular assessment throughout the entire duration of the treatment that lasted 2-3 years. The next three groups show the TNS©-PV intensity at that particular time point. Since patients with an overall medium TNS©-PV intensity were not considered in this study, the number of such patients is zero in the first group. Some HN patients had medium TNS©-PV intensity (TNS©-PV greater than 3 but lesser than 8) at some points during the treatment, as seen in the next three groups.

### Longitudinal versus independent analysis of metabolite profiles

Following the collection of patient blood samples, we performed metabolite profiling using liquid chromatography with tandem mass spectrometry (LC-MS/MS) (refer to Methods for description). We then analyzed the metabolite profile across the three time points, initially using hierarchical clustering. The hierarchical clustering dendogram showed that the profiles clustered according to the time points, as shown in Figure 2. Each branch in the figure corresponds to a sample. Since the samples were clearly demarcated according to the time points, it implies that the metabolite profiles were distinctly expressed at the three time points. Following the hierarchical clustering, we used an algorithm to develop a longitudinal support vector classifier (SVC) [36] model to further confirm the distinct expression of metabolites at the three time points. The algorithm was used to estimate a parameter *β* according to the following equation:

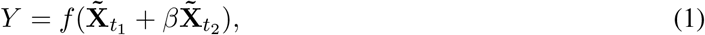

where, *t*_1_ and *t*_2_ are two time points, *Y* is the neuropathy response, and 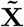 is the metabolite expression matrix. This algorithm was used to estimate *β* by minimizing the error between the predicted and actual response. For different combinations of the time points, *β* was approximately zero, regardless of the initial guess, implying that no identical set of metabolites can accurately predict neuropathy at different time points. Based on these two results, we hypothesized that different metabolites should be predictive of neuropathy at the different time points leading us to analyze the profiles separately as discussed in the next section.

**Figure 2:**
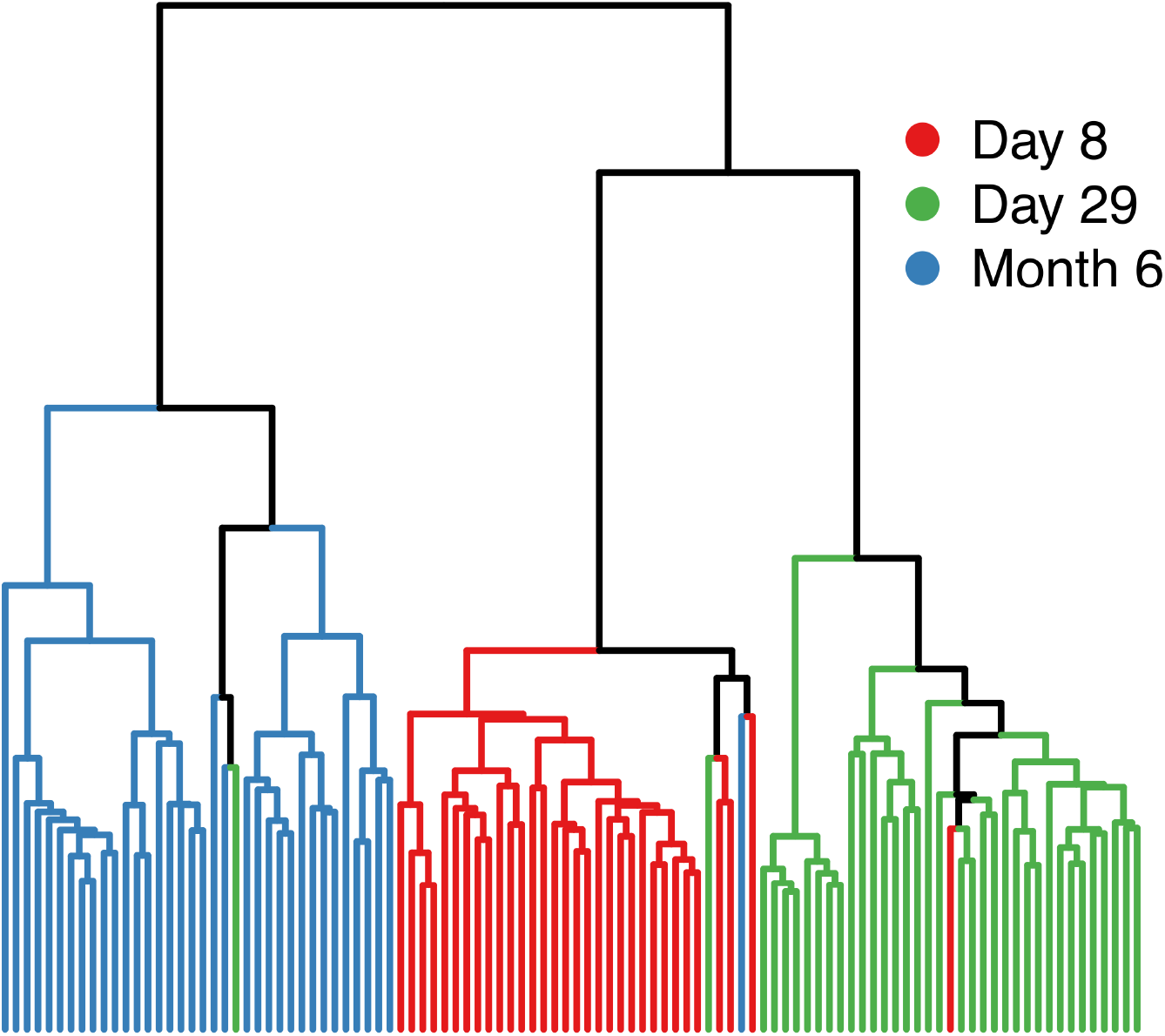
A dendogram created based on the Euclidean distance shows that the metabolite profiles are clustered according to their corresponding time points. Day 8 and day 29 metabolite profiles belong to the same primary branch and are consequently closer to each other.

### Metabolite selection and model building

In order to find a small set of metabolites that can accurately predict overall susceptibility to neuropathy, we used recursive feature elimination (RFE) along with cross validation to develop a linear SVC model. Since hierarchical clustering and longitudinal SVC modeling (described in the previous section) indicated that the metabolite profiles are distinctly expressed across the three time points, we applied RFE separately on each of the time point-specific metabolite profile matrix. Our matrix of features included metabolite expression and vincristine concentration at the three time points: 5 metabolites at day 8, 46 metabolites at day 29, and 42 metabolites at the month 6 time point were chosen, with an area under the receiver operating characteristics curve (AUROC) of 0.97, 0.95, and 0.96, respectively. The vincristine concentration was not one of the selected features at any time point. As shown in Table 2A, these selected metabolites performed well at predicting neuropathy at all the three time points, with an average AUROC greater than 0.90 and standard deviation of AUROC less than 0.1.

**Table 2:** Metrics obtained by performing RFE on the data sets at the three time points. A: The set of metabolites found that can accurately predict overall neuropathy susceptibility (HN versus LN) at these time points before manual integration of the chromatogram peaks. B: The set of metabolites found that can accurately predict TNS©-PV intensity of either high or low at that specific time point. C: The set of metabolites that can accurately predict overall neuropathy susceptibility at the time points after manual integration of peaks. AUROC: Area Under Receiver Operating Characteristics Curve, AUROCSD: standard deviations for AUROC. See Supplementary Table S1 for sensitivity and specificity corresponding to each of these.

|  | Time point | Predictors | AUROC | AUROCSD |
| --- | --- | --- | --- | --- |
| A | Day 8 | 5 | 0.968 | 0.048 |
|  | Day 29 | 46 | 0.946 | 0.060 |
|  | Month 6 | 42 | 0.963 | 0.043 |
| B | Day 29 | 2 | 0.831 | 0.120 |
|  | Month 6 | 1955 | 0.812 | 0.086 |
| C | Day 8 | 6 | 0.938 | 0.047 |
|  | Day 29 | 48 | 0.861 | 0.122 |
|  | Month 6 | 45 | 0.923 | 0.069 |

We also performed RFE to find metabolites that can predict time point-specific neuropathy (i.e., high or low intensity of TNS©-PV) in patients (Table 2B). Since only 1 patient had high TNS©-PV during day 8, we did not perform the analysis on that data set. From the day 29 metabolite profile data, only 2 metabolites were chosen that could predict TNS©-PV intensity at that point, with an AUROC of 0.83 (lower accuracy than that obtained from the previous analysis shown in Table 2A). From the month 6 data, a small set of metabolites could not be selected; 1955 metabolites were chosen (Table 2B) with a lower accuracy (AUROC 0.81) as compared to the previous case (Table 2A). This implies that the metabolites are more effective in classifying patients based on the overall VIPN susceptibility, rather than VIPN intensity at the specific time points.

After selecting the small set of metabolites, that can classify between HN and LN, using RFE, we investigated integration of their chromatogram peaks at the corresponding retention times to ensure that they were correctly integrated and were not chosen because of any potential error in integration. In order to rigorously validate the selected metabolites as biomarkers, we manually reviewed integration of peaks at the retention times of every selected metabolite, for every sample, and at all the time points. We used the Agilent ProFinder software for this purpose. We found the peaks by matching the m/z and retention time from the metabolite profile matrix to the data loaded in ProFinder and then manually visualized each of the peaks. If any integration was not accurate, we re-integrated that peak. This resulted in a polished metabolite profile matrix.

We repeated the RFE with this polished metabolite profile matrix to find metabolites that can classify between HN and LN. Using this polished data, 6 metabolites were selected at day 8, 48 metabolites at day 29, and 45 metabolites at month 6 time point, with AUROC of 0.94, 0.86, and 0.92, respectively. Table 2C shows the metrics obtained from the model fitting. In this case, the day 8 and month 6 metabolites performed better than those from the day 29 data, based on the AUROC and corresponding standard deviation.

To obtain the final list of accurately integrated and predictive metabolites, we chose metabolites that were selected both from the polished (2C) and the unpolished data (2A) using RFE on each of them. We further discarded the metabolites whose peaks looked erroneous and were difficult to properly integrate. This refined procedure provided a final list of 2, 14, and 21 metabolites from day 8, day 29, and month 6 data, respectively. We performed repeated cross validation again to train an SVC model using these final set of metabolites. Repeated cross validation was performed to estimate the predictability of the model and to find the optimal tuning parameter (the “cost” parameter) for the SVC model. The average cross validation AUROC was 0.93, 0.75, and 0.91 for day 8, day 29, and month 6 data, respectively. The average ROC’s for the trained models are shown in Figure 3.

**Figure 3:**
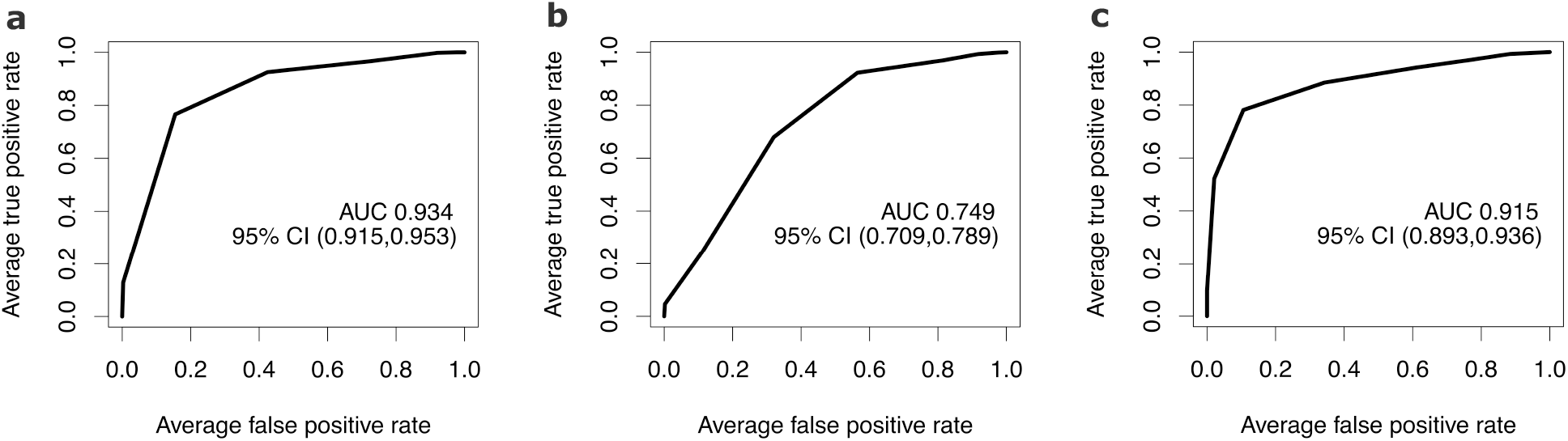
ROC plots for the final trained models at the three time points. a: Day 8, b: Day 29, c: Month 6. AUC: Area Under Curve. CI: Confidence Interval.

After training the model corresponding to each time point using repeated cross validation, we chose an optimal probability threshold to classify patients based on the output from the SVC model. To find the optimal probability threshold, we evaluated the cross validation trained models’ accuracy upon varying the threshold value. Evaluation of an appropriate threshold is needed since our data is unbalanced; keeping a threshold of 0.5 will lead to a bias towards HN. Supplementary Figure S1 shows sensitivity, specificity, Youden’s J statistic, and distance from best possible cutoff (i.e. sensitivity and specificity equal to 1) for the day 8, day 29, and month 6 trained models, where the models were built with the optimal tuning parameters obtained from cross validation. We chose the threshold based on the minimum of distance from the best possible cutoff which led to thresholds of 0.7, 0.65, and 0.7 for the three respective time points. We finally evaluated the cross validated trained models’ performance based on this newly determined threshold. Model performance was based on a confusion matrix (provided in Supplementary Table S2) and metrics calculated as shown in Table 3. The metrics shown in Table 3 show the ability of the models to make predictions when trained using a subset of the data. We have only reported cross validation accuracy metrics since we did not have a test data set to assess the performance of our final trained model. We finally trained the model using the whole data set and the optimal tuning parameters, which can be used to test new patient data.

**Table 3:** Cross validation accuracy metrics for the models with optimal tuning using the final selected metabolites. These metrics were calculated after choosing the probability thresholds for each of the time points. Cost = 4, 0.25, 0.25 for day 8, day 29, and month 6 models, respectively.

| Model Metric | Day 8 | Day 29 | Month 6 |
| --- | --- | --- | --- |
| Accuracy | 0.842 | 0.753 | 0.815 |
| 95% CI | (0.812, 0.870) | (0.720, 0.784) | (0.785, 0.843) |
| No Information Rate (NIR) | 0.750 | 0.667 | 0.667 |
| P-Value [Accuracy > NIR] | 1.07e-08 | 3.14e-07 | < 2.20e-16 |
| Kappa | 0.6093 | 0.448 | 0.625 |
| McNemar’s Test P-Value | 3.41e-4 | 0.600 | < 2.20e-16 |
| Sensitivity | 0.856 | 0.806 | 0.754 |
| Specificity | 0.800 | 0.646 | 0.938 |
| Positive Predictive Value | 0.928 | 0.820 | 0.960 |
| Negative Predictive Value | 0.650 | 0.625 | 0.656 |
| Prevalence | 0.750 | 0.667 | 0.667 |
| Detection Rate | 0.642 | 0.538 | 0.503 |
| Detection Prevalence | 0.692 | 0.656 | 0.524 |
| Balanced Accuracy | 0.828 | 0.726 | 0.846 |

We used multiple metrics to determine the accuracy and validity of the predictive models. First, the p value of the one sided test for an accuracy greater than the No Information Rate is less than 0.05 for all the three time point models. This implies that the models predict better than simply random guessing. However, with accuracy as a metric, day 8 and month 6 data outperform. The balanced accuracy ((sensitivity + specificity)/2) is higher for day 8 and 6 month data as well, indicating that the model for day 8 and month 6 data seems to be more reliable in predicting overall VIPN susceptibility. We have developed a user-friendly interface, named *VIPNp*, to use our trained models for day 8 and month 6 time points, freely available at GitHub [34] and the Shinyapps server [33].

### Metabolite structure identification

After we obtained the final list of 2, 14, and 21 metabolites at day 8, day 29, and month 6, respectively, we wanted to identify the molecular structure of these metabolites based on their mass-to-charge ratio (m/z), retention times, and tandem mass spectrometry (MS/MS) spectra. MS/MS spectra was available for only a subset of the final list: 1 metabolite from the day 8 list, 5 metabolites from the day 29 list, and 8 metabolites from the month 6 list. MS/MS spectra was compared to the METLIN [37] database and the Human Metabolome Database (HMDB) [38]. No match was found for the day 8 metabolite; for day 29, one metabolite matched with Adenosine 5’-monophosphate (AMP) for all the collision energies; for month 6 data, one metabolite matched with L-pipecolic acid for one of the collision energies. In order to confirm the identity of these metabolites, MS/MS of commercially sourced AMP and pipecolic acid standards was performed to determine the retention times. The retention times of the external compounds matched with that from the patient data (1 min of commercially sourced pipecolic acid versus 0.802 min as identified from the data, 1.4 min versus 1.77 min for AMP) confirming the identity of these two metabolites.

We used the LC-MS CEU Mass Mediator (CMM) search tool [39, 40] with m/z value, adduct, and retention time inputs in the HMDB database to identify remaining metabolites. The most reasonable options were shortlisted. For example, if one of the searches was a metabolite that could only be found due to consumption of alcohol, it was discarded as a potential option since our focus in this study is on pediatric patients. Except for 1,7-Dimethylguanosine and Phenylalanylproline, all other potentially identified metabolites were at least previously detected in blood, according to HMDB. Using these approaches, we were able to identify 4 and 9 metabolites in the day 29 and month 6 lists, respectively. Tables 4 and 5 define the identified metabolites from the day 29 and month 6 lists (Supplementary Tables S3, S4 and S5 contain the complete list of metabolites, their mass, retention time, and adduct information).

**Table 4:** Identified metabolites that can accurately predict neuropathy susceptibility at the day 29 time point.

| HMDB ID | Name | KEGG ID |
| --- | --- | --- |
| HMDB0003357 | N-Acetylornithine | C00437 |
| HMDB0000757 | Glycogen | C00182 |
| HMDB0000045 | Adenosine monophosphate | C00020 |
| HMDB0001341 | Adenosine diphosphate | C00008 |

**Table 5:** Identified metabolites that can accurately predict neuropathy susceptibility at the month 6 time point.

| HMDB ID | Name | KEGG ID |
| --- | --- | --- |
| HMDB0000716 | L-Pipecolic acid | C00408 |
| HMDB0013464 | SM(d18:0/16:1(9Z)) | C00550 |
| HMDB0000670 | Homo-L-arginine | C01924 |
| HMDB0001961 | 1,7-Dimethylguanosine |  |
| HMDB0010383 | LysoPC(16:1(9Z)) | C04230 |
| HMDB0003337 | Oxidized glutathione | C00127 |
| HMDB0011170 | gamma-Glutamylisoleucine |  |
| HMDB0001855 | 5-Hydroxytryptophol |  |
| HMDB0011177 | Phenylalanylproline |  |

### Pathway analysis

In order to investigate the relevance of the chosen and identified metabolites, we used the pathway analysis tool in Metaboanalyst 4.0 [41, 42]. We selected the hypergeometric test for over representation analysis and the relative-betweeness centrality for pathway topology analysis.

For day 29, two metabolites (Adenosine diphosphate, Adenosine monophosphate) were identified to be part of purine metabolism, and one metabolite (N-Acetylornithine) was identified to be part of the arginine biosynthesis (details in Supplementary Table S6). Since there were very limited metabolites to perform pathway analysis and only one or two of them belonged to a specific pathway which originally consisted of several metabolites, none of the metabolite to pathway associations were statistically significant after adjustment of the p values required to account for multiple testing. Furthermore, we did not find evidence for the role of these pathways in chemotherapy-induced peripheral neuropathy in the existing literature.

For the set of month 6 metabolites, SM(d18:0/16:1(9Z)), Oxidized glutathione, LysoPC(18:1(9Z)), and Pipecolic acid were identified to be part of the sphingolipid metabolism, glutathione metabolism, glycerophospholipid metabolism, and lysine degradation, respectively (details in Supplementary Table S7). Again, none of the pathway and metabolite associations were statistically significant after adjustment of the p values to account for multiple testing. We, however, did find evidence for the role of sphingolipid metabolism in chemotherapy-induced peripheral neuropathy [43–45]. Moreover, glutathione is a popular antioxidant tested as a therapeutic for chemotherapy-induced peripheral neuropathy, with various related studies mentioned in a review by Starobova and Vetter [46]. There is also evidence for the involvement of glycerophospholipid metabolism [47] which indicates that these metabolites may be biologically relevant to VIPN.

## Discussion

Peripheral neuropathy is a painful and debilitating side-effect of vincristine, a common chemotherapeutic drug used for treatment of pediatric ALL patients, as well as, many other pediatric and adult cancers. Currently, there is no established way of predicting VIPN during the initial stage of the treatment. Identification of specific biomarkers will aid in adjusting the vincristine dose according to susceptibility of patients to VIPN in order to improve their quality of life. Even though all previous omics related studies have focused on genomics, it is imperative to include phenotypic variabilities to find VIPN predictors given their known impact on drug response. In this study, we investigated the role of metabolites in predicting susceptibility of ALL pediatric patients to VIPN.

We performed metabolite profiling of ALL patients at three time points during the treatment: day 8, day 29, and month 6. First, we found that the metabolite profiles were distinctly expressed at the three time points, as shown by hierarchical clustering (Figure 2) and longitudinal SVC modeling, indicating that the overall metabolite profile varied as a direct response to the treatment. Second, preliminary analysis using RFE resulted in sets of 5, 46, and 42 metabolites that accurately predicted overall VIPN susceptibility. Since the vincristine concentration was not one of the selected features from RFE, we concluded that regardless of the vincristine concentration at the time points, a small set of metabolites can accurately predict overall neuropathy susceptibility in these patients at these time points. Third, we manually integrated chromatogram peaks of these selected metabolites and performed RFE again with the polished matrix. Subsequently, we chose the metabolites that overlapped from the analysis with unpolished and polished data. Then, final sets of 2, 14, and 21 metabolites were found that could predict overall VIPN susceptibility at day 8, day 29, and month 6, respectively.

Metabolites could not predict time point-specific TNS©-PV intensity as accurately as overall susceptibility to VIPN (LN versus HN). This further strengthened our hypothesis that specific downstream metabolites can be potential biomarkers of overall neuropathy susceptibility due to treatment with vincristine. Furthermore, the final models built with the chosen metabolites were more accurate in predicting neuropathy at day 8 and month 6 as compared to the model built for the day 29 data (Table 3), implying that our developed predictive models can be used to evaluate VIPN susceptibility at the day 8 and month 6 time points during the treatment.

Based on the final developed models at the day 8 and month 6 time points, we present a framework for predicting a patient’s overall VIPN susceptibility during ALL treatment (Figure 4). On day 8 of the treatment, blood samples can be collected for metabolite profiling, specifically to obtain expression of the 2 chosen metabolites. The expression of these metabolites can be used to predict overall neuropathy susceptibility from our trained model. If the output of the model is a probability greater than the threshold value of 0.7, the physician may need to lower the vincristine dose as that sample might correspond to an HN patient. A follow up evaluation can be performed after 6 months by obtaining expression of the 21 chosen metabolites (Supplementary Table S5). If the output from the two trained models is consistently a probability greater than 0.7, the physician could consider lowering the vincristine dose. This framework can aid in vincristine dose decision making for the physicians. Our user-friendly tool, *VIPNp* [34, 33], can be used to execute this strategy.

**Figure 4:**
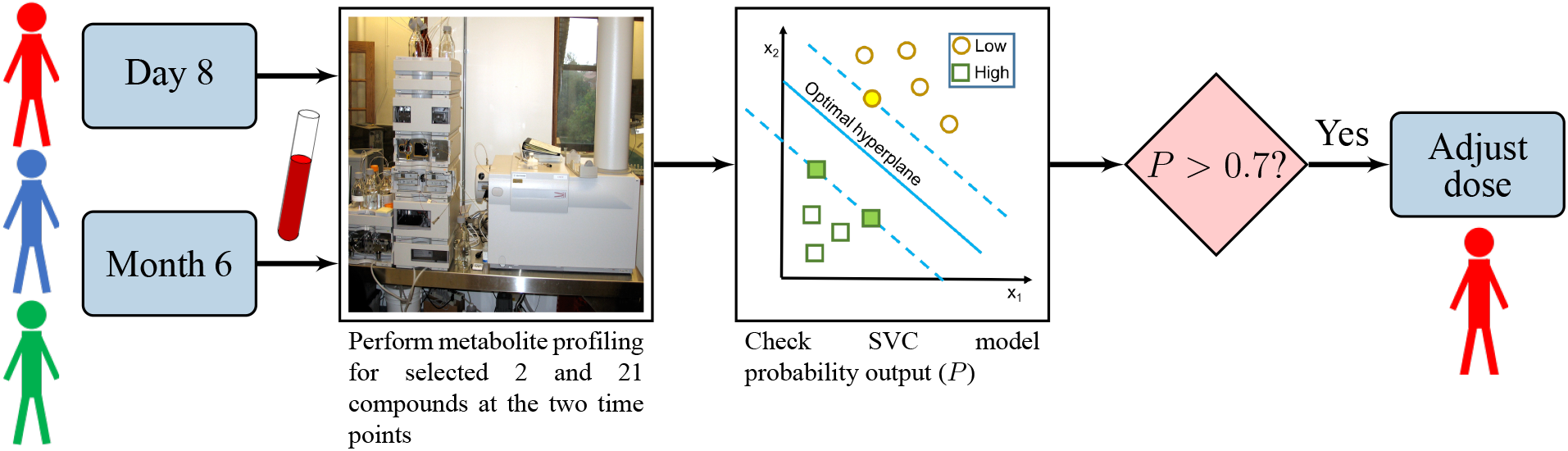
A workflow showing a potential vincristine dose decision making strategy based on the trained SVC models. Blood samples of patients can be collected at day 8 and month 6 time points of the treatment. Samples can then be analyzed using mass spectrometry for metabolite profiling of the selected 2 and 21 metabolites at the day 8 and month 6 time points, respectively. The metabolite profile data can then be used to predict overall neuropathy susceptibility from our trained SVC models. If the model output probability is greater than a threshold value of 0.7, the patient might be susceptible to overall high neuropathy (HN). This strategy enables identification of patients susceptible to HN. The vincristine dose for HN patients may require adjustment.

In the foregoing suggested strategy, a significant caveat is that these models can only classify patients as susceptible to HN or LN. Some patients might be susceptible to “medium” VIPN (TNS©-PV greater than 3 but lesser than 8) during the treatment. Such cases have not been included here, implying that the models have limited predictability and that they should be used with caution. As a result, we only suggest that if the output of the model is greater than 0.7 at both day 8 and month 6 predictions, then there may be a need to adjust vincristine dose, since such patients are most likely classified as HN.

Even though we finalized a list of metabolites using RFE (Supplementary Tables S3, S4, and S5), we could not identify all of them. This greatly reduced our capability to find the biological relevance of these chosen metabolites. We could not obtain MS/MS spectra for all of them, which may be a result of their presence in smaller quantities. Even after obtaining the MS/MS spectra, we could not find matches from existing databases for most of these metabolites. As a result, we only used the available m/z value, retention time, and adduct information for the remaining unidentified metabolites, which does not ensure that we have identified the metabolites correctly. For a thorough identification, MS/MS spectra is needed for all of the chosen metabolites and subsequently the retention times of those compounds need to be validated using authentic standards. Since only a few metabolites could be identified and consequently used for pathway analysis, none of the associations between the metabolites and their pathways found were statistically significant. Even though some of the identified metabolites for the month 6 data belonged to pathways that are relevant to chemotherapy-induced peripheral neuropathy, their biological relevance could not be established with certainty.

The peak integration of chromatograms also needs to be interpreted with caution. We observed that some peaks may not be integrated correctly or may simply be difficult to integrate for only a few metabolites. This has two implications: 1) The chosen metabolites may be predictive of neuropathy because they were improperly integrated. We eliminated this possibility by manually integrating and repeating RFE with a polished metabolite profile matrix. 2) Some potential biomarker metabolites might not have been chosen because of poor integration. It is not feasible to manually inspect every peak for every sample given that there were approximately 2000 metabolites and 104 samples (including all the time points), leaving a possibility that all potential predictors might not have been identified. Regardless, the current metabolites found at day 8 and month 6 are certainly potential predictors since they accurately predict VIPN susceptibility.

It should also be noted that the metabolite profile matrix is defined as high dimensional–with several metabolites and very few samples. This leaves a possibility that, despite using feature selection machine learning algorithms, some potential metabolites may have been overlooked. Moreover, we do not have balanced samples; having an equal number of high and low neuropathy patients will allow for a more accurate model. We also do not have a test set to validate our model. It is critical to perform this study with more samples and additional datasets in order to test and validate our model. *VIPNp* [34, 33] can be used to test our trained models on new datasets in the future. There is also a need to include “medium” neuropathy patients to accurately predict multiple outcomes of the treatment. Since this is the first metabolomics based pilot study for VIPN, we only focused on HN and LN patients to explore the potential of this approach in predicting VIPN susceptibility.

Despite these limitations, our preliminary study has shown that ALL treatment can alter the metabolite profiles, and a few selected metabolites can accurately predict the overall VIPN susceptibility. We have also provided a strategy (Figure 4) to adjust the vincristine dose based on VIPN susceptibility at two time points: day 8 and month 6. Our work shows that metabolomics can aid in predicting VIPN susceptibility during the early stage of treatment. Although it must be admitted that our prediction of high and low neuropathy could have had a stronger statistical backing with a larger cohort of patients, we contend that the numbers in this study were not so unreasonably small as to challenge this major conclusion. Based on an exhaustive literature review, all previous studies have only focused on genomics and pharmacokinetics in the prediction of chemotherapy-induced peripheral neuropathy with the exception of one proteomics study [48]. This pilot study is the first of its kind focused on metabolomics to predict VIPN. In order for this methodology to be more effective, we need a balanced and large number of fasting samples, accurate integration of chromatogram peaks, MS/MS spectra for all the chosen metabolites, and a more exhaustive database to identify these metabolites.

Peripheral neuropathy is not just limited to vincristine but is a dose-limiting toxicity of several other chemotherapy drugs (paclitaxel, taxane, and cisplatin), as well. A problem of this scale requires expertise and continued collaboration of individuals across multiple disciplines. An integrative approach involving better quantitation of all the omics data, large patient cohorts, careful phenotyping of patient data, and state of the art machine learning and statistical techniques is necessary in order to find a robust prediction of VIPN and any other chemotherapy-induced peripheral neuropathy, even before the start of the treatment.

## Methods

### ALL patient data collection [49]

Children with newly diagnosed precursor B-cell ALL were recruited from four academic medical centers: Indiana University School of Medicine/Riley Hospital for Children, the University of Michigan Comprehensive Cancer Center/Mott Children’s Hospital, Vanderbilt University Medical Center/Monroe Carell Jr. Children’s Hospital, and George Washington University/Children’s National Medical Center. Participants were between the ages of 1 and 18 at the time of diagnosis and received vincristine according to Pediatric Oncology Group (POG) treatment trials (including: POG studies 9904 and 9905). The standard vincristine dosage received was 1.5 mg/m^2^ (capped at 2-mg maximum dose). Toxicity-based dose modifications were defined according to the specific POG protocol guiding the individual child’s leukemia treatment. Patients were excluded if they had any of the following criteria: baseline peripheral neuropathy score greater than grade 1 per the NCI CTCAE©version 4.0; currently receiving erythropoietin, itraconazole, or vitamin supplement greater than 100% of the recommended daily allowance; Down’s syndrome; pregnancy; or a history of coexisting serious illness that would limit neurological assessments. All procedures were reviewed by the Indiana University Internal Review Board and approved (protocol number 1105005420). All the methods were performed in accordance with the relevant guidelines and regulations. Informed consent was obtained from a parent and/or legal guardian for study participation.

### Metabolomics sample preparation and extraction

Protein removal and sample extraction were performed by adding 200 *µ*L methanol to 100 *µ*L of plasma. Solutions were sonicated for 2 minutes, chilled at −20 °C for 1 hour, and centrifuged at 16,000 *×* g for 8 minutes. The supernatants were transferred to separate vials and evaporated to dryness in a vacuum concentrator. The dried fractions were reconstituted in 75 *µ*L of diluent composed of 95% water and 5% acetonitrile, containing 0.1% formic acid. Solutions were sonicated for 5 minutes, centrifuged at 16,000 *×* g for 8 minutes, and the supernatants were transferred to plastic HPLC total recovery autosampler vials.

### HPLC-MS analysis of metabolomics samples [50]

Separations were performed on an Agilent 1290 system (Palo Alto, CA), with a mobile phase flow rate of 0.45 mL/min. The metabolites were assayed using a Waters HSS T3 UPLC column (1.8 *µ*m, 2.1 × 100 mm), where the mobile phase A and B were 0.1% formic acid in ddH_2_O and acetonitrile, respectively. Initial conditions were 100:0 A:B, held for 1 minute, followed by a linear gradient to 70:30 at 16 min, then 5:95 at 21 min. Column re-equilibration was performed by returning to 100:0 A:B at 22 minutes and holding until 27 minutes. The mass analysis was obtained using an Agilent 6545 Q-TOF mass spectrometer with electrospray ionization (ESI) capillary voltage +3.2 kV, nitrogen gas temperature 325 °C, drying gas flow rate 8.0 L/min, nebulizer gas pressure 30 psig, fragmentor voltage 130 V, skimmer 45 V, and OCT RF 750 V. Mass data from m/z 70-1000 scans were collected at 5 Hz using Agilent MassHunter Acquisition software (v. B.06). Mass accuracy was improved by infusing Agilent Reference Mass Correction Solution (G1969-85001). To assist with compound identification, MS/MS was performed in a Data-dependent Acquisition mode. Five precursors per cycle were obtained using fixed collision energies of 10, 20, and 40 eV.

Peak deconvolution, integration, and alignment was performed using Agilent ProFinder (v. B.06). Peak annotations were performed using the METLIN (www.metlin.scripps.edu) and HMDB (www.hmdb.ca) metabolite databases, with a mass error of less than 15 ppm.

### Vincristine quantitation [51]

#### Sample preparation

100 *µ*L plasma was pipetted to a microcentrifuge tube. Protein precipitation was performed by adding 200 *µ*L cold methanol. The mixture was vortexed for 2 minutes, chilled at −20 °C for 1 hour, and then centrifuged at 13,000 g for 8 minutes. Supernatant was transferred to a fresh microcentrifuge tube and vacuum concentrated to dryness. Reconstituted in 75 *µ*L of diluent (95% water and 5% acetonitrile, with 0.1% formic acid), sonicated for 5 minutes, centrifuged at 13,000 rpm for 8 minutes, and supernatant was transferred to HPLC vials.

#### HPLC/MS-MS analysis

Vincristine plasma levels were quantitated by HPLC/MS-MS. Separation was performed on an Agilent Rapid Res 1200 HPLC system using an Agilent Zorbax XDB-C18 (2.1 *×* 50 mm, 3.5 *µ*m) column. Mobile phase A was water with 0.1% formic acid and mobile phase B was acetonitrile with 0.1% formic acid. Initial conditions were 95:5 A:B, held for 0.5 minute, followed by a linear gradient to 0:100 at 8 min, and held until 10 min. Column re-equilibration was performed by returning to 95:5 A:B at 11 minutes and held until 15 minutes. Column flow rate was 0.3 mL/min. Retention time for vincristine was 5.9 minutes.

Analytes were quantified by MS/MS utilizing an Agilent 6460 triple quadrupole mass spectrometer with electrospray ionization (ESI). Quantitation was based on Multiple Reaction Monitoring (MRM). ESI positive mode was used with a transition of 825.2 to 807.7 (quantifier) and 825.2 to 765.3 (qualifier), with a collision energy (CE) of 45 and 40 V, respectively. A fragmentor energy of 135 V and a dwell time of 80 ms was used. Source parameters were as follows: nitrogen gas temperature = 330 °C and flow rate = 10 L/min, nebulizer pressure = 35 psi, sheath gas temperature = 250 °C, sheath gas flow rate = 7 L/min, and capillary potential = 3.5 kV. All the data were collected and analyzed with Agilent MassHunter software. Quantitation was based on a 6 point standard curve, with concentration range from 0.1 to 500 ng/mL, by spiking vincristine into unmedicated human plasma (Sigma). Standard curves were fit to a quadratic function, with a 1/x curve fit weighting. Correlation coefficients *>* 0.99 were obtained.

### Metabolite profiling data analysis

Data analysis was performed in R [52]. First, metabolites that were not present in at least 75% of the patients in at least one group were discarded. Subsequently, missing data imputation was performed using a modified k-nearest neighbours (KNN) imputation as specified in [53]. Impute package in R was used to perform the KNN imputation [54]. Hierarchical clustering was performed using hclust, with ward.D2 as the method and Euclidean distance as the metric. For metabolite selection, recursive feature elimination (RFE) along with repeated 5-fold cross validation support vector classifier was used. Repetition was done 20 times. This was further iterated 100 times and the model was chosen based on highest AUROC and lowest specificity standard deviation. For recursive feature elimination, all sizes of number of predictors between 1 to 50 were allowed. Caret [55] package from R was used for this. The method specified in caret for this algorithm is svmLinear2, from the package e1071 [56]. Tunelength was kept as 1 and hence the cost parameter was fixed to 0.25. SVM with linear kernel was chosen because more tuned parameters would be needed for nonlinear kernels. Since our dataset had few samples, that may have lead to overfitting. Final model fitting results specified in Table 2 were based on resampling from bootstrapping 25 times, using the selected number of predictors. This was the default model fitting mode in caret while performing RFE.

For model training after finalizing the set of metabolites, we trained using 5-fold repeated cross validation, repeated 20 times. The metric for accuracy was kept as AUROC. Here, tuneLength was kept as 10. Cost function of the SVM model was tuned using this. After model training, for deciding the probability threshold, thresholds between 0.5 and 1 were explored. The one corresponding to minimum distance from perfect sensitivity and specificity (i.e. both of them equal to 1) was chosen. Subsequently, confusion matrix and its metrics were generated using caret package’s confusion matrix generator.

## Data Availability

Our trained models are available on https://github.com/parulv1/VIPNp. The metabolite profiles are available from the corresponding author on reasonable request.

## Acknowledgements

This project was funded, in part, with support from Grant Number R01HD062484-06 from the National Institutes of Health and National Institute of Child Health and Human Development, and Indiana Clinical and Translational Sciences Institute funded, in part by Grant Number UL1TR002529 from the National Institutes of Health, National Center for Advancing Translational Sciences, Clinical and Translational Sciences Award. The content is solely the responsibility of the authors and does not necessarily represent the official views of the National Institutes of Health. P.V. would like to thank Lina Aboulmouna for reviewing the manuscript and Kanishka Misra for helping with developing *VIPNp*.

## Author contributions statement

J.S., T.S., E.S., R.H., R.H., E.W., L.L., and J.R. collected the samples and phenotyped the patients. B.C. performed metabolite profiling and vincristine quantitation. P.V., J.D., J.S., J.R., B.C, and D.R. designed the study. P.V. generated and analyzed the results. P.V, T.S., B.C., and D.R. wrote the manuscript. P.V., J.S., B.C., and D.R. reviewed the manuscript.

## Additional information

The authors declare no competing interests.

## Notes

### Competing Interest Statement

The authors have declared no competing interest.

## References

[1] Elizabeth Ward, Carol DeSantis, Anthony Robbins, Betsy Kohler, and Ahmedin Jemal. Childhood and adolescent cancer statistics, 2014. CA: a cancer journal for clinicians, 64(2):83–103, 2014.

[2] American Cancer Society, Cancer Facts & Figures 2018. https://www.cancer.org/research/cancer-facts-statistics/all-cancer-facts-figures/cancer-facts-figures-2018.html. Accessed: 04-08-2019.

[3] Erika Mora, Ellen M Lavoie Smith, Clare Donohoe, and Daniel L Hertz. Vincristine-induced peripheral neuropathy in pediatric cancer patients. American journal of cancer research, 6(11):2416, 2016.

[4] Mirjam E. van de Velde, Gertjan L. Kaspers, Floor C.H. Abbink, Abraham J. Wilhelm, Johannes C.F. Ket, and Marleen H. van den Berg. Vincristine-induced peripheral neuropathy in children with cancer: A systematic review. Critical reviews in oncology/hematology, 114:114–130, 2017.

[5] Ellen M Lavoie Smith, Lang Li, ChienWei Chiang, Karin Thomas, Raymond J Hutchinson, Elizabeth M Wells, Richard H Ho, Jodi Skiles, Arindom Chakraborty, Celia M Bridges, et al. Patterns and severity of vincristine-induced peripheral neuropathy in children with acute lymphoblastic leukemia. Journal of the Peripheral Nervous System, 20(1):37–46, 2015.

[6] Satu S Lehtinen, Usko E Huuskonen, Arja H Harila-Saari, Uolevi Tolonen, Leena K Vainionpää, and B Marjatta Lanning. Motor nervous system impairment persists in long-term survivors of childhood acute lymphoblastic leukemia. Cancer: Interdisciplinary International Journal of the American Cancer Society, 94(9):2466–2473, 2002.

[7] Kirsten K Ness, Melissa M Hudson, Ching-Hon Pui, Daniel M Green, Kevin R Krull, Tseng T Huang, Leslie L Robison, and E Brannon Morris. Neuromuscular impairments in adult survivors of childhood acute lymphoblastic leukemia: associations with physical performance and chemotherapy doses. Cancer, 118(3):828–838, 2012.

[8] Heleen A Reinders-Messelink, Marina M Schoemaker, Marjorie Hofte, Ludwig NH Göeken, Annet Kingma, Meta M van den Briel, and Willem A Kamps. Fine motor and handwriting problems after treatment for childhood acute lymphoblastic leukemia. Medical and Pediatric Oncology, 27(6):551–555, 1996.

[9] Cinzia R De Luca, Maria McCarthy, Jane Galvin, Jessica L Green, Alexandra Murphy, Sarah Knight, and Jacqueline Williams. Gross and fine motor skills in children treated for acute lymphoblastic leukaemia. Developmental neurorehabilitation, 16(3):180–187, 2013.

[10] Kirsten K Ness, Ann C Mertens, Melissa M Hudson, Melanie M Wall, Wendy M Leisenring, Kevin C Oeffinger, Charles A Sklar, Leslie L Robison, and James G Gurney. Limitations on physical performance and daily activities among long-term survivors of childhood cancer. Annals of internal medicine, 143(9):639–647, 2005.

[11] Doralina L. Anghelescu, Lane G. Faughnan, Sima Jeha, Mary V. Relling, Pamela S. Hinds, John T. Sandlund, Cheng Cheng, Deqing Pei, Gisele Hankins, Jennifer L. Pauley, and Ching-Hon Pui. Neuropathic pain during treatment for childhood acute lymphoblastic leukemia. Pediatric Blood & Cancer, 57(7):1147–1153, 2011.

[12] Barthelemy Diouf, Kristine R Crews, Glen Lew, Deqing Pei, Cheng Cheng, Ju Bao, Jie J Zheng, Wenjian Yang, Yiping Fan, Heather E Wheeler, et al. Association of an inherited genetic variant with vincristine-related peripheral neuropathy in children with acute lymphoblastic leukemia. Jama, 313(8):815–823, 2015.

[13] Shinji Kishi, Cheng Cheng, Deborah French, Deqing Pei, Soma Das, Edwin H. Cook, Nobuko Hijiya, Carmelo Rizzari, Gary L. Rosner, Tony Frudakis, Ching-Hon Pui, William E. Evans, and Mary V. Relling. Ancestry and pharmacogenetics of antileukemic drug toxicity. Blood, 109(10):4151–4157, 2007.

[14] Jamie L Renbarger, Kevin C McCammack, Caroline E Rouse, and Stephen D Hall. Effect of race on vincristine-associated neurotoxicity in pediatric acute lymphoblastic leukemia patients. Pediatric blood & cancer, 50(4):769–771, 2008.

[15] Andrew S Moore, Ross Norris, Gareth Price, Thu Nguyen, Ming Ni, Rani George, Karin van Breda, John Duley, Bruce Charles, and Ross Pinkerton. Vincristine pharmacodynamics and pharmacogenetics in children with cancer: A limited-sampling, population modelling approach. Journal of Paediatrics and Child Health, 47(12):875–882, 2011.

[16] Richard Aplenc, Wendy Glatfelter, Peggy Han, Eric Rappaport, Mei La, Avital Cnaan, M Anne Blackwood, Beverly Lange, and Timothy Rebbeck. CYP3A genotypes and treatment response in paediatric acute lymphoblastic leukaemia. British journal of haematology, 122(2):240–244, 2003.

[17] Akinbode Egbelakin, Michael J. Ferguson, Emily A. MacGill, Amalia S. Lehmann, Ariel R. Topletz, Sara K. Quinney, Lang Li, Kevin C. McCammack, Stephen D. Hall, and Jamie L. Renbarger. Increased risk of vincristine neurotoxicity associated with low CYP3A5 expression genotype in children with acute lymphoblastic leukemia. Pediatric Blood & Cancer, 56(3):361–367, 2011.

[18] Romain Guilhaumou, Caroline Solas, Veronique Bourgarel-Rey, Sylvie Quaranta, Angelique Rome, Nicolas Simon, Bruno Lacarelle, and Nicolas Andre. Impact of plasma and intracellular exposure and CYP3A4, CYP3A5, and ABCB1 genetic polymorphisms on vincristine-induced neurotoxicity. Cancer chemotherapy and pharmacology, 68(6):1633–1638, 2011.

[19] Francesco Ceppi, ChloéLanglois-Pelletier, Vincent Gagné, Julie Rousseau, Claire Ciolino, Samanta De Lorenzo, Kojok M Kevin, Diana Cijov, Stephen E Sallan, Lewis B Silverman, Donna Neuberg, Jeffery L Kutok, Daniel Sinnett, Caroline Laverdiére, and Maja Krajinovic. Polymorphisms of the vincristine pathway and response to treatment in children with childhood acute lymphoblastic leukemia. Pharmacogenomics, 15(8):1105–1116, 2014. PMID: 25084203.

[20] Elixabet Lopez-Lopez, Angela Gutierrez-Camino, Itziar Astigarraga, Aurora Navajas, Aizpea Echebarria-Barona, Purificacion Garcia-Miguel, Nagore Garcia de Andoin, Carmen Lobo, Isabel Guerra-Merino, Idoia Martin-Guerrero, and Africa Garcia-Orad. Vincristine pharmacokinetics pathway and neurotoxicity during early phases of treatment in pediatric acute lymphoblastic leukemia. Pharmacogenomics, 17(7):731–741, 2016. PMID: 27180762.

[21] Ellis Groninger, Tiny Meeuwsen-de Boer, Pauline Koopmans, Donald Uges, Wim Sluiter, Anjo Veerman, Willem Kamps, and Siebold de Graaf. Pharmacokinetics of vincristine monotherapy in childhood acute lymphoblastic leukemia. Pediatric Research, 52(1):113, 2002.

[22] William R Crom, Siebold SN de Graaf, Timothy Synold, Donald RA Uges, Henk Bloemhof, Gaston Rivera, Michael L Christensen, Hazem Mahmoud, and William E Evans. Pharmacokinetics of vincristine in children and adolescents with acute lymphocytic leukemia. The Journal of pediatrics, 125(4):642–649, 1994.

[23] CEM Gidding, GJ Meeuwsen-de Boer, P Koopmans, DRA Uges, WA Kamps, and SSN De Graaf. Vincristine pharmacokinetics after repetitive dosing in children. Cancer chemotherapy and pharmacology, 44(3):203–209, 1999.

[24] HW Van den Berg, ZR Desai, R Wilson, G Kennedy, JM Bridges, and RG Shanks. The pharmacokinetics of vincristine in man. Cancer chemotherapy and pharmacology, 8(2):215–219, 1982.

[25] CEM Gidding, SJ Kellie, WA Kamps, and SSN De Graaf. Vincristine revisited. Critical reviews in oncology/hematology, 29(3):267–287, 1999.

[26] W Stock, B Diouf, KR Crews, D Pei, C Cheng, K Laumann, SJ Mandrekar, S Luger, A Advani, RM Stone, RA Larson, and WE Evans. An Inherited Genetic Variant in CEP72 Promoter Predisposes to Vincristine-Induced Peripheral Neuropathy in Adults With Acute Lymphoblastic Leukemia. Clinical Pharmacology & Therapeutics, 101(3):391–395, 2017.

[27] Galen E.B. Wright, Ursula Amstutz, Britt I. Drögemöller, Joanne Shih, Shahrad R. Rassekh, Michael R. Hayden, Bruce C. Carleton, Colin J.D. Ross, and The Canadian Pharmacogenomics Network for Drug Safety Consortium. Pharmacogenomics of Vincristine-Induced Peripheral Neuropathy Implicates Pharmacokinetic and Inherited Neuropathy Genes. Clinical Pharmacology & Therapeutics, 105(2):402–410, 2019.

[28] Angela Gutierrez-Camino, Idoia Martin-Guerrero, Elixabet Lopez-Lopez, Aizpea Echebarria-Barona, Iñaki Zabalza, Irune Ruiz, Isabel Guerra-Merino, and Africa Garcia-Orad. Lack of association of the CEP72 rs924607 TT genotype with vincristine-related peripheral neuropathy during the early phase of pediatric acute lymphoblastic leukemia treatment in a Spanish population. Pharmacogenetics and genomics, 26(2):100–102, 2016.

[29] Barthelemy Diouf and William E. Evans. Pharmacogenomics of Vincristine-Induced Peripheral Neuropathy: Progress Continues. Clinical Pharmacology & Therapeutics, 105(2):315–317, 2019.

[30] D Jayachandran, Usha Ramkrishna, Jodi Skiles, Jamie Renbarger, and Doraiswami Ramkrishna. Revitalizing personalized medicine: respecting biomolecular complexities beyond gene expression. CPT: pharmacometrics & systems pharmacology, 3(4):1–11, 2014.

[31] R Kaddurah-Daouk, R Weinshilboum, and on behalf of the Pharmacometabolomics Research Network. Metabolomic Signatures for Drug Response Phenotypes: Pharmacometabolomics Enables Precision Medicine. Clinical Pharmacology & Therapeutics, 98(1):71–75, 2015.

[32] Bingbing Li, Xuyun He, Wei Jia, and Houkai Li. Novel Applications of Metabolomics in Personalized Medicine: A Mini-Review. Molecules, 22(7), 2017.

[33] VIPNp. https://parulv1.shinyapps.io/vipnp_shiny/.

[34] VIPNp. https://github.com/parulv1/VIPNp.

[35] Ellen M Lavoie Smith, Lang Li, Raymond J Hutchinson, Richard Ho, W Bryan Burnette, Elizabeth Wells, Ms Celia Bridges, and Jamie Renbarger. Measuring vincristine-induced peripheral neuropathy in children with acute lymphoblastic leukemia. Cancer nursing, 36(5):E49, 2013.

[36] Shuo Chen and F DuBois Bowman. A novel support vector classifier for longitudinal high-dimensional data and its application to neuroimaging data. Statistical Analysis and Data Mining: The ASA Data Science Journal, 4(6):604–611, 2011.

[37] Colin A Smith, Grace O’Maille, Elizabeth J Want, Chuan Qin, Sunia A Trauger, Theodore R Brandon, Darlene E Custodio, Ruben Abagyan, and Gary Siuzdak. METLIN: a metabolite mass spectral database. Therapeutic drug monitoring, 27(6):747–751, 2005.

[38] David S Wishart, Yannick Djoumbou Feunang, Ana Marcu, An Chi Guo, Kevin Liang, Rosa Vázquez-Fresno, Tanvir Sajed, Daniel Johnson, Carin Li, Naama Karu, Zinat Sayeeda, Elvis Lo, Nazanin Assempour, Mark Berjanskii, Sandeep Singhal, David Arndt, Yonjie Liang, Hasan Badran, Jason Grant, Arnau Serra-Cayuela, Yifeng Liu, Rupa Mandal, Vanessa Neveu, Allison Pon, Craig Knox, Michael Wilson, Claudine Manach, and Augustin Scalbert. H MDB 4.0: the human metabolome database for 2018. Nucleic Acids Research, 46(D1):D608–D617, 11 2017.

[39] Alberto Gil de la Fuente, Joanna Godzien, Mariano Fernández López, Francisco J Rupérez, Coral Barbas, and Abraham Otero. Knowledge-based metabolite annotation tool: CEU Mass Mediator. Journal of pharmaceutical and biomedical analysis, 154:138–149, 2018.

[40] Alberto Gil-de-la Fuente, Joanna Godzien, Sergio Saugar, Rodrigo Garcia-Carmona, Hasan Badran, David S Wishart, Coral Barbas, and Abraham Otero. CEU Mass Mediator 3.0: a metabolite annotation tool. Journal of proteome research, 18(2):797–802, 2018.

[41] Jasmine Chong and Jianguo Xia. MetaboAnalystR: an R package for flexible and reproducible analysis of metabolomics data. Bioinformatics, 34(24):4313–4314, 2018.

[42] Jasmine Chong, Othman Soufan, Carin Li, Iurie Caraus, Shuzhao Li, Guillaume Bourque, David S Wishart, and Jianguo Xia. MetaboAnalyst 4.0: towards more transparent and integrative metabolomics analysis. Nucleic acids research, 46(W1):W486–W494, 2018.

[43] Kali Janes, Joshua W Little, Chao Li, Leesa Bryant, Collin Chen, Zhoumou Chen, Krzysztof Kamocki, Timothy Doyle, Ashley Snider, Emanuela Esposito, et al. The development and maintenance of paclitaxel-induced neuropathic pain require activation of the sphingosine 1-phosphate receptor sub-type 1. Journal of Biological Chemistry, 289(30):21082–21097, 2014.

[44] Rita Kramer, Jacek Bielawski, Emily Kistner-Griffin, Alaa Othman, Irina Alecu, Daniela Ernst, Drew Kornhauser, Thorsten Hornemann, and Stefka Spassieva. Neurotoxic 1-deoxysphingolipids and paclitaxel-induced peripheral neuropathy. The FASEB Journal, 29(11):4461–4472, 2015.

[45] Katherine Stockstill, Timothy M Doyle, Xisheng Yan, Zhoumou Chen, Kali Janes, Joshua W Little, Kathryn Braden, Filomena Lauro, Luigino Antonio Giancotti, Caron Mitsue Harada, et al. Dysregulation of sphingolipid metabolism contributes to bortezomib-induced neuropathic pain. Journal of Experimental Medicine, 215(5):1301–1313, 2018.

[46] Hana Starobova and Irina Vetter. Pathophysiology of chemotherapy-induced peripheral neuropathy. Frontiers in molecular neuroscience, 10:174, 2017.

[47] Fei-ze Wu, Wen-juan Xu, Bo Deng, Si-da Liu, Chao Deng, Meng-yu Wu, Yu Gao, and Li-qun Jia. Wen-Luo-Tong Decoction Attenuates Paclitaxel-Induced Peripheral Neuropathy by Regulating Linoleic Acid and Glycerophospholipid Metabolism Pathways. Frontiers in pharmacology, 9, 2018.

[48] Emily I. Chen, Katherine D. Crew, Meghna Trivedi, Danielle Awad, Mathew Maurer, Kevin Kalinsky, Antonius Koller, Purvi Patel, Jenny Kim Kim, and Dawn L. Hershman. Identifying Predictors of Taxane-Induced Peripheral Neuropathy Using Mass Spectrometry-Based Proteomics Technology. PLOS ONE, 10(12):1–15, 12 2016.

[49] Lang Li, Tammy Sajdyk, Ellen M. L. Smith, Chien Wei Chang, Claire Li, Richard H. Ho, Raymond Hutchinson, Elizabeth Wells, Jodi L. Skiles, Naomi Winick, Paul L. Martin, and Jamie L. Renbarger. Genetic variants associated with vincristine-induced peripheral neuropathy in two populations of children with acute lymphoblastic leukemia. Clinical Pharmacology & Therapeutics, 105(6):1421–1428, 2019.

[50] Sherleen Xue-Fu Adamson, Ruoxing Wang, Wenzhuo Wu, Bruce Cooper, and Jonathan Shannahan. Metabolomic insights of macrophage responses to graphene nanoplatelets: Role of scavenger receptor cd36. PLOS ONE, 13(11):1–30, 11 2018.

[51] Fen Yang, Hongyun Wang, Ming Liu, Pei Hu, and Ji Jiang. Determination of free and total vincristine in human plasma after intravenous administration of vincristine sulfate liposome injection using ultrahigh performance liquid chromatography tandem mass spectrometry. Journal of Chromatography A, 1275:61 –69, 2013.

[52] R Core Team. R: A Language and Environment for Statistical Computing. R Foundation for Statistical Computing, Vienna, Austria, 2017.

[53] Emily Grace Armitage, Joanna Godzien, Vanesa Alonso-Herranz, Ángeles López-Gonzálvez, and Coral Barbas. Missing value imputation strategies for metabolomics data. Electrophoresis, 36(24):3050–3060, 2015.

[54] Trevor Hastie, Robert Tibshirani, Balasubramanian Narasimhan, and Gilbert Chu. impute: Imputation for microarray data, 2017. R package version 1.50.1.

[55] Max Kuhn. Contributions from Jed Wing, Steve Weston, Andre Williams, Chris Keefer, Allan Engelhardt, Tony Cooper, Zachary Mayer, Brenton Kenkel, the R Core Team, Michael Benesty, Reynald Lescarbeau, Andrew Ziem, Luca Scrucca, Yuan Tang, Can Candan, and Tyler Hunt. caret: Classification and Regression Training. R package version 6.0-78.

[56] David Meyer, Evgenia Dimitriadou, Kurt Hornik, Andreas Weingessel, and Friedrich Leisch. e1071: Misc Functions of the Department of Statistics, Probability Theory Group (Formerly: E1071), TU Wien, 2017. R package version 1.6-8.

